# Assessing Quantitative Performance and Expert Review of Multiple Deep Learning-Based Frameworks for Computed Tomography-based Abdominal Organ Auto-Segmentation

**DOI:** 10.1101/2024.10.02.24312658

**Authors:** Udbhav S. Ram, Joel A. Pogue, Michael Soike, Neil T. Pfister, Rojymon Jacob, Carlos E. Cardenas

## Abstract

Segmentation of abdominal organs in clinical oncological workflows is crucial for ensuring effective treatment planning and follow-up. However, manually generated segmentations are time-consuming and labor-intensive in addition to experiencing inter-observer variability. Many deep learning (DL) and Automated Machine Learning (AutoML) frameworks have emerged as a solution to this challenge and show promise in clinical workflows. This study presents a comprehensive evaluation of existing AutoML frameworks (Auto3DSeg, nnU-Net) against a state-of-the-art non-AutoML framework, the Shifted Window U-Net Transformer (SwinUNETR), each trained on the same 122 training images, taken from the Abdominal Multi-Organ Segmentation (AMOS) grand challenge. Frameworks were compared using Dice Similarity Coefficient (DSC), Surface DSC (sDSC) and 95th Percentile Hausdorff Distances (HD95) on an additional 72 holdout-validation images. The perceived clinical viability of 30 auto-contoured test cases were assessed by three physicians in a blinded evaluation. Comparisons show significantly better performance by AutoML methods. nnU-Net (average DSC: 0.924, average sDSC: 0.938, average HD95: 4.26, median Likert: 4.57), Auto3DSeg (average DSC: 0.902, average sDSC: 0.919, average HD95: 8.76, median Likert: 4.49), and SwinUNETR (average DSC: 0.837, average sDSC: 0.844, average HD95: 13.93). AutoML frameworks were quantitatively preferred (13/13 OARs p>0.0.5 in DSC and sDSC, 12/13 OARs p>0.05 in HD95, comparing Auto3DSeg to SwinUNETR, and all OARs p>0.05 in all metrics comparing SwinUNETR to nnU-Net). Qualitatively, nnU-Net was preferred over Auto3DSeg (p=0.0027). The findings suggest that AutoML frameworks offer a significant advantage in the segmentation of abdominal organs, and underscores the potential of AutoML methods to enhance the efficiency of oncological workflows.

## Introduction

This study aims to address the impactful challenge of automating abdominal organ segmentation in CT images, a task that is critical for robust radiotherapy planning workflows and which traditionally relies on time-consuming processes, which are also usually subject to inter-observer variability. While increasingly automated machine and deep learning related advances have been leveraged to alleviate these concerns, there remains significant questions in the areas of which of the latest models/frameworks offer the largest gains in performance and efficiency and whether there may be any significantly better alternatives to ‘manual’ deep learning methodologies compared to AutoML solutions. Furthermore, the evaluation of segmentations generated by deep learning methods lack real world, qualitative analysis by physicians and rather rely on only qualitative metrics. By conducting a comprehensive analysis of non-AutoML vs. AutoML frameworks, and looking at AutoML to AutoML comparisons, this study aims to highlight particular shortcomings with more manual methods, taking into account physician preference. Recently, several reports (Cardenas et al.^1^, Yu et al.^2^, Wong et al.^3^) have shown the promise of deep learning-based techniques as a possible tool in advancing clinical auto-segmentation workflows. Adding to this, Chung et al.^4^ conducted a comprehensive Turing test where participants showed a preference for deep learning-based auto-segmentations over manual segmentations, showing the clinical validity and impact of choosing a potential model to train and deploy for any prospective clinical workflows. In recent years, ‘automatic machine learning,’ or AutoML for short, has gained in popularity as it has reduced the need to develop training pipelines and pre/post-processing workflows. AutoML techniques have been successfully applied to auto segmentation tasks in the form of various frameworks such as the nnU-Net and Auto3DSeg. Isensee et al.^5^ introduced the nnU-Net in 2019 as an AutoML framework that works by combining state-of-the-art deep learning frameworks with dataset-specific data augmentation, and a user-friendly framework into an open-source project. nnU-Net has been a forerunner in AutoML applications in biomedical image segmentation and has shown to be robust, adaptable, and precise at many internationally recognized auto-segmentation competitions, including the AMOS22 Grand Challenge (Isensee et al.^6^). As such, nnU-Net, is considered a baseline approach to benchmark new auto-segmentation methods. Several works have been published in the years since its release detailing its utility in a variety of clinical segmentation workflows^7^,^8^. nnU-Net has had many applications, not limited to organ segmentation, but even in anomaly detection and segmentation of tumors. Shu et al.^9^ showed the efficacy of nnU-Net auto-segmenting pituitary adenomas, and others (Pettit et al.^10^) have shown robust results in liver volume determination and parenchyma. Moreover, work done by Yu et al.^2^ and Ziyaee et al.^11^ have shown the advantages of the nnU-Net in abdominal and brain metastases segmentation, respectively.

More recently, the Auto3DSeg framework was open sourced through the Medical Open Network for Artificial Intelligence (MONAI) Project^12^. Auto3DSeg presents some similarities to nnU-Net in that both frameworks require minimal user input. Furthermore, Auto3DSeg leverages a holistic evaluation of the entire dataset and statistical analysis to determine a training structure and assorted hyper-parameters. While the two frameworks are similar in their operation, Auto3DSeg has the added advantage of increased transparency, modularity, and ease of modification for any specific application. This added ability to modify the AutoML process itself to fit a researcher’s needs promises additional gains in performance and accuracy. Since its introduction, Auto3DSeg has captivated the medical computer vision community and has been touted as a potential competing framework to the nnU-Net. While Auto3DSeg has only been recently released, works by Siddiquee et al.^13^, have shown its efficacy in 3D segmentation workflows for identification of intracranial hemorrhages. Furthermore, the head and neck tumor segmentation and outcome prediction (HECKTOR) challenge report contained several groups (He et al.^14^, Myronenko et al.^15^) who leveraged the Auto3DSeg framework to develop effective auto-segmentation models, which further confirmed the competitive nature of the Auto3DSeg pipeline when compared to existing state-of-the-art model training frameworks.

The present study shows novel comparisons and evaluations of these frameworks, particularly in the following areas of interest: (1) **Comparison of manual and AutoML frameworks**: This study aims to conduct a first-of-its-kind, comprehensive head-to-head comparison of two leading AutoML frameworks, nnU-Net and Auto3DSeg. (2) **Using publicly available data** To improve the reproducibility and transparency of the work conducted, we used data which is publicly available from the AMOS22 Grand Challenge, and systematically evaluate both models on their ability to perform abdominal organ segmentation in computed tomography (CT) images. (3) **Quantitative and qualitative evaluation**: We provide a detailed quantitative analysis based on Dice Similarity Coefficient (DSC), Surface DSC (sDSC), and 95th percentile Hausdorff Distances (HD95). Moreover, we incorporate a qualitative assessment by expert physicians through blinded evaluations of 30 auto-segmented test cases, offering clinical insights into the usability and preference of the generated segmentations. We hypothesize that the differences in auto-segmentation outcomes between these frameworks are minimal. Moreover, both AutoML approaches are expected to significantly outperform traditional non-AutoML segmentation methods in terms of accuracy and efficiency.

## Materials and Methods

### Data Description

In this study, we utilized the CT data from the publicly available Abdominal Multi-Organ Segmentation 2022 dataset^16^ (from the MICCAI AMOS22 Grand Challenge), a diverse, multi-modality cohort containing 500 CT and 100 MRI scans, each provided with radiotherapy structure sets (The Dataset is available for download here: https://zenodo.org/records/7155725#.Y0OOCOxBztM). Each individual CT scan contains voxel level annotations of 15 target organs. The 15 provided organs are: spleen, right and left kidneys, gall bladder, esophagus, stomach, aorta, postcava, right and left adrenal glands, duodenum, bladder, and prostate or uterus (depending on the sex). The publishers of the AMOS22 dataset released training data (200 CT scans) and unannotated testing data (100 CT scans) in the first phase of evaluation. Here, we set aside these 100 cases without labels for qualitative evaluation (see model evaluation). For the 200 training CT scans, 78 scans were set aside for final quantitative evaluation as a holdout-validation set. Out of these 78 cases, 72 were chosen to conduct statistical and quantitative analysis on, with the selection criteria being any scan containing 13 ground-truth OARs. The 6 removed images presented cases with missing OARs due to prostatectomy/hysterectomy, cholecystectomy, or any OAR not being visible in the field of view of the scan. The remaining 122 training images underwent a 5-fold cross-validation process, where cross-validation splits were held consistent across different frameworks to minimize training cohort bias in head-to-head comparison. Prior to training, bilateral organs (i.e., left kidney/right kidney and left adrenal/right adrenal) were combined into a single organ to take advantage of left-right flip augmentations.

### Frameworks

We aimed to benchmark three distinct training frameworks in this study, those being, a standard SwinUNETR framework (non-AutoML) proposed by Hatamizadeh et al.^17^, the nnU-Net framework (AutoML) and the MONAI Auto3DSeg framework (AutoML). The U-Net, introduced by Ronneberger et al.^18^, is a convolutional neural network architecture that aims to capture image context using a contracted-expanding path layout (giving it its distinctive U-like shape), which enables precise localization and forms the basis for the SwinUNETR and nnU-Net as well as certain models trialed within Auto3DSeg.

#### nnU-Net

The nnU-Net is trained using a in a three-step process. The first step is defining training parameters; here, we use default parameters such as the loss function, data augmentation, and learning rate, which were defined during nnU-Net’s initial development. Following this, the next step determines individual dataset’s fingerprint, which optimize patch size, network topology and batch size within the computational constraints of the available GPU. Lastly, the nnU-Net preprocesses training data to allow training for three architectures: 2D U-Net, 3D, Full Resolution U-Net (3D U-Net) and a 3D, Cascaded U-Net (3DC U-Net). Each architecture is trained using a 5-fold cross-validation approach. After all folds and architectures are trained, the nnU-Net provides a mechanism to determine optimal architecture ensembling and segmentation post-processing. Finally, the ensembled model is used for inference on the pre-defined and unseen test data. A simplified visualization of the nnU-Net pipeline is illustrated in Figure 1 (b).

**Figure 1.**
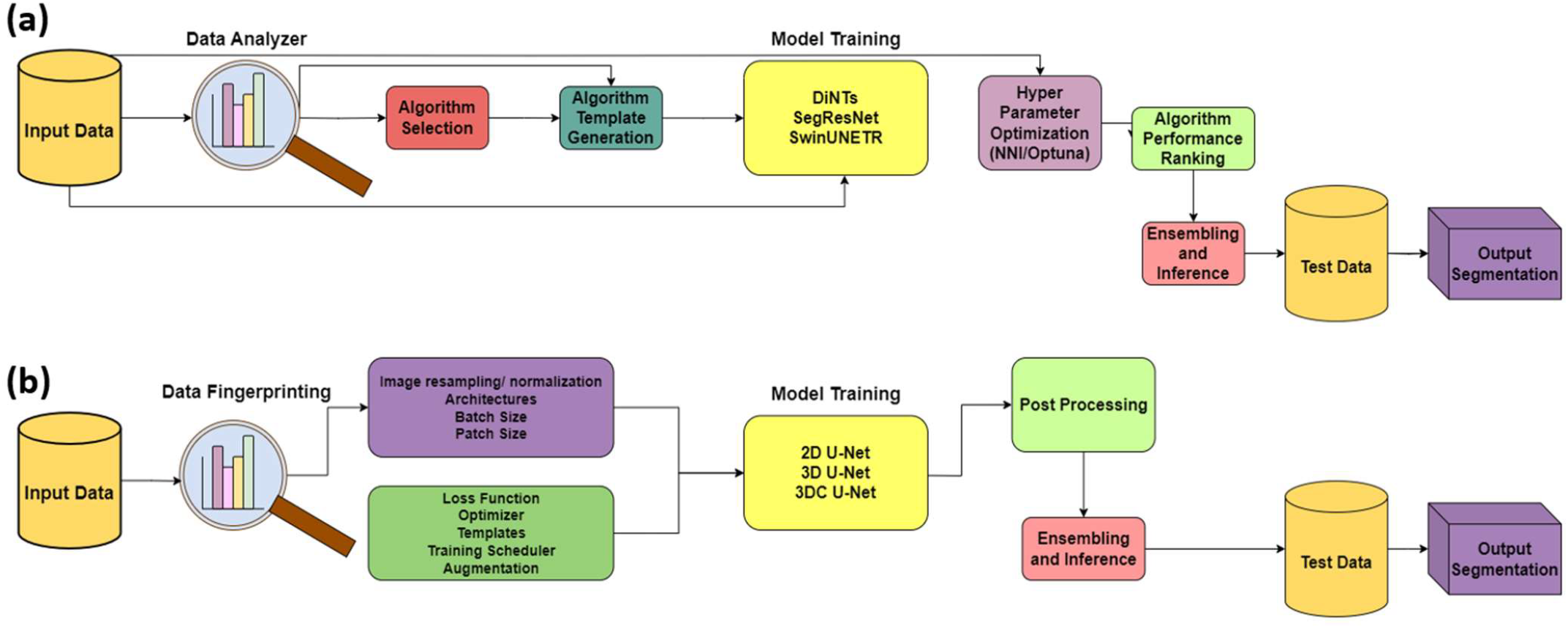
Comparing both AutoML pipelines (a): Auto3DSeg, (b): nnU-Net

#### Auto3DSeg

Similar to the nnU-Net framework, Auto3DSeg aims to deliver a user-friendly application to train deep learning segmentation models. Once a list of data and the location of the data is provided, Auto3DSeg implements a training pipeline, which consists of the steps shown in Figure 1 (a). During data analysis, the data analyzer examines a given medical image dataset and reports the statistics of image intensity, shape, spacing, etc. This is analogous to the ‘data fingerprinting’ approach used by the nnU-Net. For the algorithm generation, the framework creates self-contained algorithm folders for model training, inference, and validation with all proposed network architectures and training recipes. The default algorithms are based on three different networks: DiNTS^19^, (2D/3D) SegResNet based on^20^ the omission of variational autoencoders, and the addition of allowing 2D input, and a SwinUNETR^17^, with their respective well-tuned training schemes. To achieve robust predictions for unseen data, the Auto3DSeg pipeline provides a model ensemble module to summarize predictions from various trained models. The module firstly ranks checkpoints of different algorithms based on validation accuracy in each fold of N-fold cross-validation, picks the top-M algorithms from each fold, and creates ensemble predictions using MN checkpoints. The default ensemble algorithm uses SoftMax/sigmoid activations to provide mean probability maps of candidate predictions and proceeds to generate the final outputs.

#### SwinUNETR

This model is a U-Net-based architecture which uses shifted window (Swin) transformers and encoders to address issues in 3D semantic segmentation tasks. The Swin model is especially useful at overcoming the limited kernel sizes of the convolutional layers in more traditional fully connected neural networks. With transformer models coming to the forefront in natural language processing and computer vision applications, the SwinUNETR represents a logical and positive change in 3D medical image segmentation. In the specific case of abdominal multi-organ segmentation, the problem is reduced to a sequence prediction, using the multi-dimensional input data and re-projecting it to a 1-Dimensional embedded sequence. These can then be used as an input to the Swin transformer and encoder pairs. Using skip connections, features can be extracted at five distinct resolutions using the shifted windows to compute the self-attention, with each resolution layer being associated with a fully connected decoder.^21^

### Pre-processing

Pre-processing in this context mainly refers to dataset augmentations applied to the original training images. While preprocessing varies for each individual framework, AutoML pre-processing is largely self-defined and framework dependent. For both approaches, an exploratory data analysis (EDA) process is employed to assess the entire dataset and generate pre-processing plans, among other parameters, that best fit the specific dataset.

For nnU-Net, EDA is defined as the ‘data fingerprinting’ step, where nnU-Net uses pre-defined, heuristic rules to infer a list of hyper-parameters, driven by the nature of the dataset. Some examples of input parameters that would influence pre/post processing strategies include the image size (pre/post crop), image spacing/voxel size, image modalities used, the total number of classes, and the number of training images. All hyperparameter spaces are captured in a .Json file that is automatically generated after the data has been fingerprinted. Data augmentation includes random combinations of rotation, scaling, gaussian noise, gaussian blur, brightness, contrast, simulation of low resolution and/or gamma resolution modifications.

With Auto3DSeg, the ‘data analyzer’ module performs a very similar task. For a given dataset, the ‘data analyzer’ reports the statistics of image intensity, shape, spacing, among other image characteristics. This analysis culminates with a summary report, known as the ‘data stats,’ which is a .yaml file containing additional, cumulative statistics such as data size, spacing, intensity distribution for the dataset, which are used to determine network characteristics and training parameters. These dataset statistics are particularly crucial in the next step of the Auto3DSeg pipeline, which is algorithm generation. Some particularly important examples of this would be how the data modality determines intensity normalization strategies, average image shape determines image region-of-interest (ROI) cropping, and input/output channels decides the first/last layers of the network.

However, when using the non-AutoML methodology, pre-processing steps are left entirely up to the user. Here, pre-processing leveraged a predefined data augmentation scheme which included resampling. The SwinUNETR training images were loaded in a specific spacing (1.5mm, 1.5mm, 2.0mm) with randomized intensity scaling to the whole image, as suggested by Tang et al. ^22^. Furthermore, the image foregrounds were cropped to help enrich the dataset by way of enhancing organs over unnecessary background information.

### Training

For each model, training was executed in a virtual machine environment on the Jetstream2 High Performance Compute cluster^23^. The g3.xl instance used to develop these models consists of an Ubuntu 22.04 environment with 200GB of disk storage, 125GB of RAM, and a GRID A100X-40C. These resources were made available through the ACCESS ecosystem^24^. Final predictions came from an ensembled model of the 5 trained folds from the winning architecture for each framework.

#### nnU-Net

All default architectures within nnU-Net, (2D U-Net, 3D U-Net and 3D-Cascaded U-Net) were trained for 1,000 epochs, with one epoch being defined as an iteration over 250 mini-batches, for a total of 250,000 iterations per model. The patch size was 40 × 263 × 263 and a batch size of 2 was used during training. Additionally, to ensure model robustness, a 5-fold cross-validation was implemented, where the 122 training images were assigned folds randomly. A combination of Dice and cross-entropy loss was used during training, as is standard in the nnU-Net pipeline.

#### Auto3DSeg

For each of these algorithms, a hyperparameter search was executed while training. In contrast to the nnU-Net framework, which required no additional modifications to the default configurations, it was necessary to modify the target re-sampling to circumvent memory issues during training. This was achieved by increasing the resolution/spacing in the transform configuration (from 1 × 1 × 1 to 2.0 × 2.0 × 2.0). An identical set of 122 training images were provided. Similar to nnU-Net, the combined Dice and cross-entropy loss was used, with an Adam optimizer and a batch size of 2.

#### SwinUNETR

The UNETR based implementation is trained for 240,000 epochs, with cross-validation every 500 epochs. The UNETR configuration was left as standard from the MONAI recommended configuration. That is, a SwinUNETR which has an image size of our standard images, typically 512 × 512 with a varying number of axial slices. We define the output channel’s parameter as the number of output classes + 1 background class, which for the combined laterality of the modified AMOS dataset, came to 14 output channels. We left the originally suggested feature size of 48. During training, Dice and cross-entropy loss was utilized using the Adam optimizer with a learning rate of 1e-4 and weight decay of 1e-5. The largest modification came from the Swin patch size, where the default 96×96×96 proved challenging in terms of resource allocation. A middle ground 64×64×64 window size was used for the final training run. There were no hyper-parameter tuning steps taken during training. Similar to the nnU-Net and Auto3DSeg training, the SwinUNETR had a 5-fold cross-validation process proceeding training with an identical split of training, testing and validation images.

### Post-Processing

nnU-Net automatically determines the postprocessing that should be used. Postprocessing in nnU-Net focuses on the removal of all components in the final prediction barring the largest, (once for foreground vs background and once for each label/volume). Similarly, Auto3DSeg develops its own post processing pipeline which is also covered entirely by the ‘autorunner’. Autorunner is the interface by which users may run the Auto3DSeg pipeline using minimal inputs. Post-processing for the non-AutoML framework largely consisted of reversing pre-processing steps to resample the resulting segmentation back to the original image space.

### Quantitative Evaluation

The resulting trained models from each framework were evaluated on the hold-out validation set of 72 scans, which contained unseen image-label pairs. These auto-segmentations were evaluated using the Dice Similarity Coefficient (DSC), 95th percentile Hausdorff distance (HD95), and Surface DSC (sDSC).

The DSC is a commonly used as an image overlap metric and it is given by the following formula, where A and B represent two volumes:

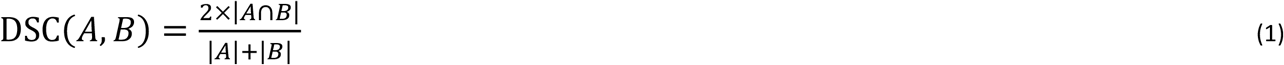

It was first proposed by Dice^25^ as a simple, normalized metric to show the overlap between two populations of interest. In the context of image segmentation, we can use the DSC to evaluate the volume of overlap between various model outputs and ground truth segmentations. In Equation 1, the DSC is given by the twice the intersection of volumes A and B, divided by the sum of the volumes. As such, the ideal ratio of 1 is attained when there is a perfect intersection of both volumes being compared, with a value of 0 representing no intersection at all. A well-trained model should aim to maximize the Dice score by achieving a score approaching one, representing complete, voxel for voxel agreement between the model output and the existing, physician generated ground truth contour.

Similarly, the Hausdorff Distance (HD) is a metric which quantifies the mutual proximity of two volumes of interest. It is given by the following formula:

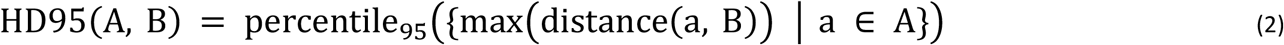

In our case, we use the 95th percentile of the set of maximum distances between the two volumes of interest (whose set of points are represented as A and B respectively, where a is the set of points belonging to volume A). The ideal scenario is an HD95 distance approaching 0mm, where 95% of points are within 0mm of each other when comparing the inferred and ground truth contours.

The final quantitative metric is the surface Dice Similarity Coefficient (sDSC), which is a relatively new metric for comparisons of volume delineations. It adds depth to the conducted quantitative analysis by quantifying deviation between organ surface contours rather than voxel-wise volumes. It was proposed by Nikolov et al.^26^ and allows for the quantification of distances between volume surfaces. Overlap is defined as the shortest distance between two surfaces falling within the pre-defined tolerance. As with the standard DSC, the sDSC can take any value from zero to one, with zero indicating no overlap and a score of one indicating complete agreement between both surfaces. A tolerance (*τ*) of 2 mm was chosen to account for a median slice spacing of 5mm across the 72 validation images.

### Qualitative Evaluation

In order to evaluate the images qualitatively, auto-contours from 30 randomly selected cases from our original 100 case test set were reviewed by three board-certified radiation oncologists with expertise treating abdominal cancers. This method of qualitative evaluation has proven to be adept for assessing potential clinical frameworks, as shown by Kraus et al.^27^ and Baroudi et al.^28^, among others. Here, the segmentations provided to the physicians were comprised of the same completely unseen testing images, which were inferenced on by all three models. The top two performing models, based on quantitative results, were selected for qualitative review. We used a randomized, blinded approach where participants did not know which structure set belonged to which model. The participants were then asked to score the auto-segmentations on a Likert scale from 1 to 5, as shown in Table 1.

**Table 1.**
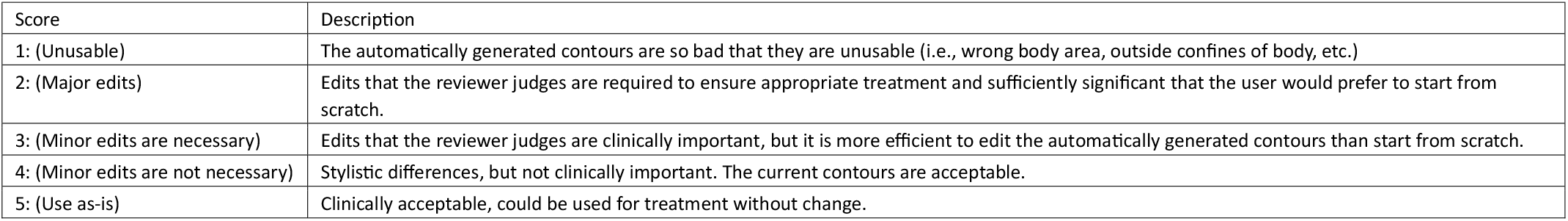
Qualitative Scoring Criteria.

### Statistical Analysis

A two-tailed Wilcoxon paired, non-parametric test was conducted on the 72-holdout images for each model pair to quantify the statistical significance of metric differences (DSC, HD95, sDSC) across all OARs, with *p*<0.05 deemed significant. Similarly, the two-sided Wilcoxon test was utilized to assess differences between physician Likert scores for the two AutoML frameworks across each OAR due to its suitability for paired, ordinal data.

A similar process is conducted on the physician generated Likert scores. The two tailed Wilcoxon signed-rank test can provide conclusive evidence of statistically significant performance differences between the 2 AutoML frameworks on any particular OARs. A statistically significant preference in one model or the other, being blinded to quantitative metrics is a viable method of assessing model outputs on a holistic and clinical level. Additionally, an unpaired t-test of distributions was conducted on all scores across both models to quantify the statistical significance of score distributions.

## Results

In this study, the ensembled 3D U-Net provided the highest performance in the nnU-Net pipeline. Similarly, Auto3DSeg cross-validation resulted in an ensembled model containing 2 folds of the 3D SegResNet and 3 folds from the SwinUNETR. The SwinUNETR only model was ensembled using the five trained folds.

### Quantitative Results

Quantitative results are summarized in Table 2, with a more detailed, per-OAR breakdown found in Figure 2. The overall summary shows a significantly better performance by both AutoML methodologies when compared to the non-AutoML UNETR framework.

**Table 2.**
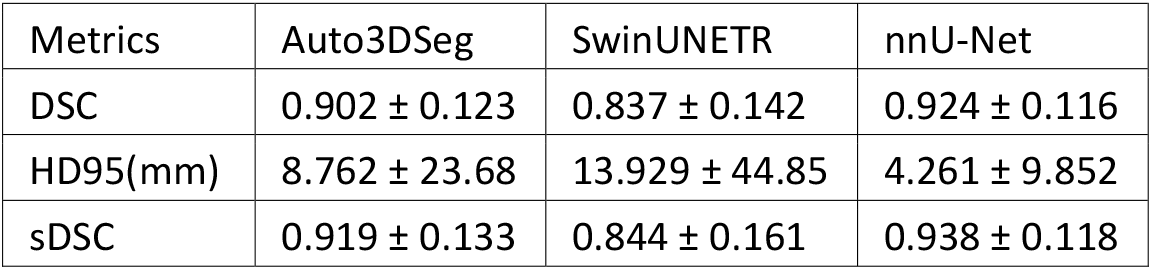
Summary of quantitative metrics for all OARs combined (average +/-standard deviations)

**Figure 2.**
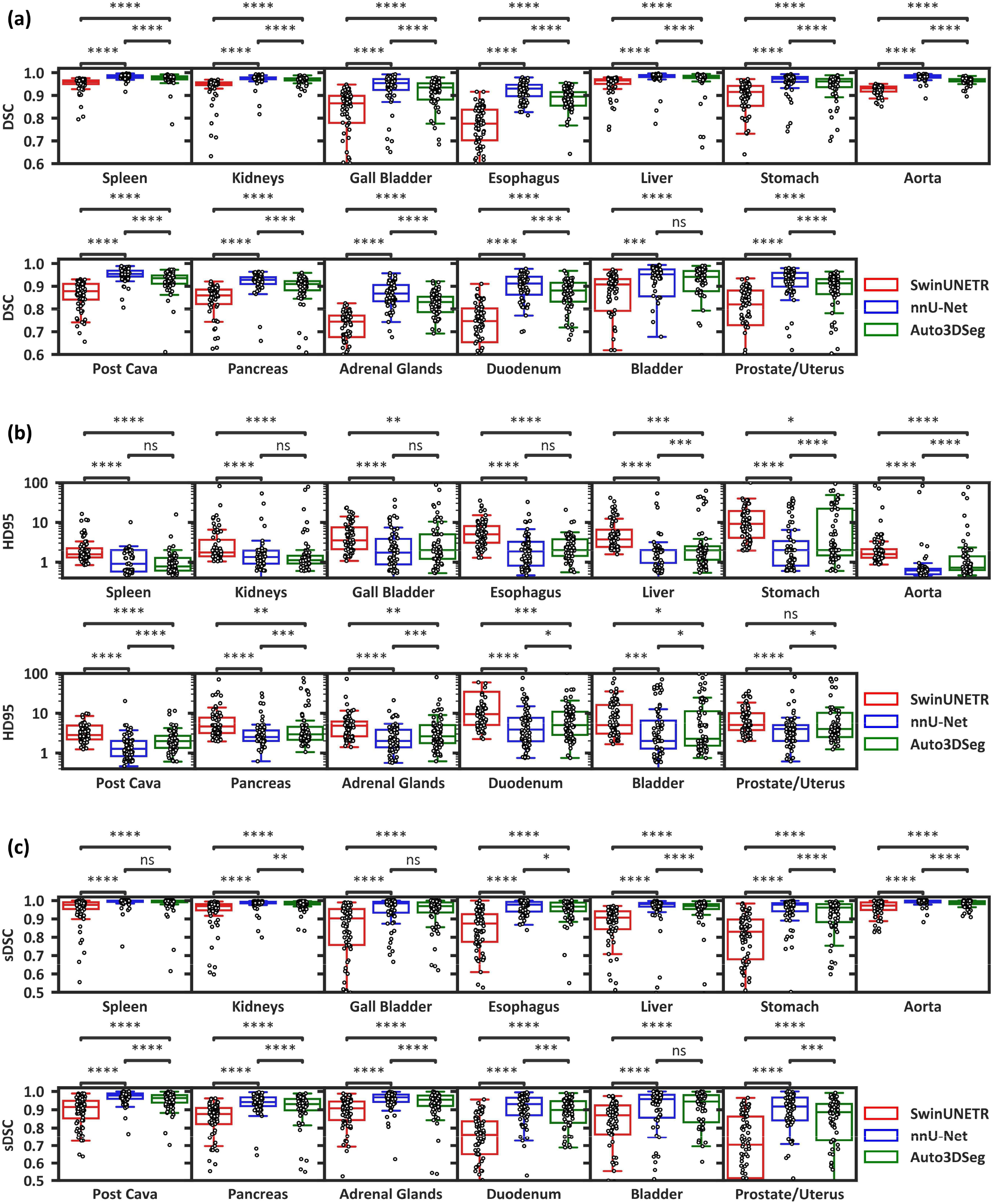
Boxplots comparing model performance across all assessed quantitative metrics (a: Dice Similarity Coefficient, b: 95th percentile Hausdorff Distance, c: Surface Dice Similarity Coefficient) for all OARs. The two-sided Wilcoxon paired non-parametric test was used for difference testing, with significance values stratified as follows: ns: p > 0.05; ^*^: 0.01 < p ≤ 0.05; ^**^: 0.001 < p ≤ 0.01; ^***^: 0.0001 < p ≤ 0.001; ^****^: p ≤ 0.0001. Ranges were selected for best visualization of the majority of data.

In general, the spleen and liver were among the highest scoring organs, across all models. Conversely, both nnU-Net and Auto3DSeg struggled with the adrenals and gall bladder OARs. For SwinUNETR, we notice a similar trend, with the liver spleen and kidneys comprising the top 3 OARs and the model being particularly challenged when contouring the adrenal glands and duodenum. All models consistently scored much lower on the Prostate/Uterus OAR. Figure 2 shows the distributions of all metrics, across all three models, for 72 holdout-validation images. An example of the model contours compared to a ground truth image is shown in Figure 3.

**Figure 3.**
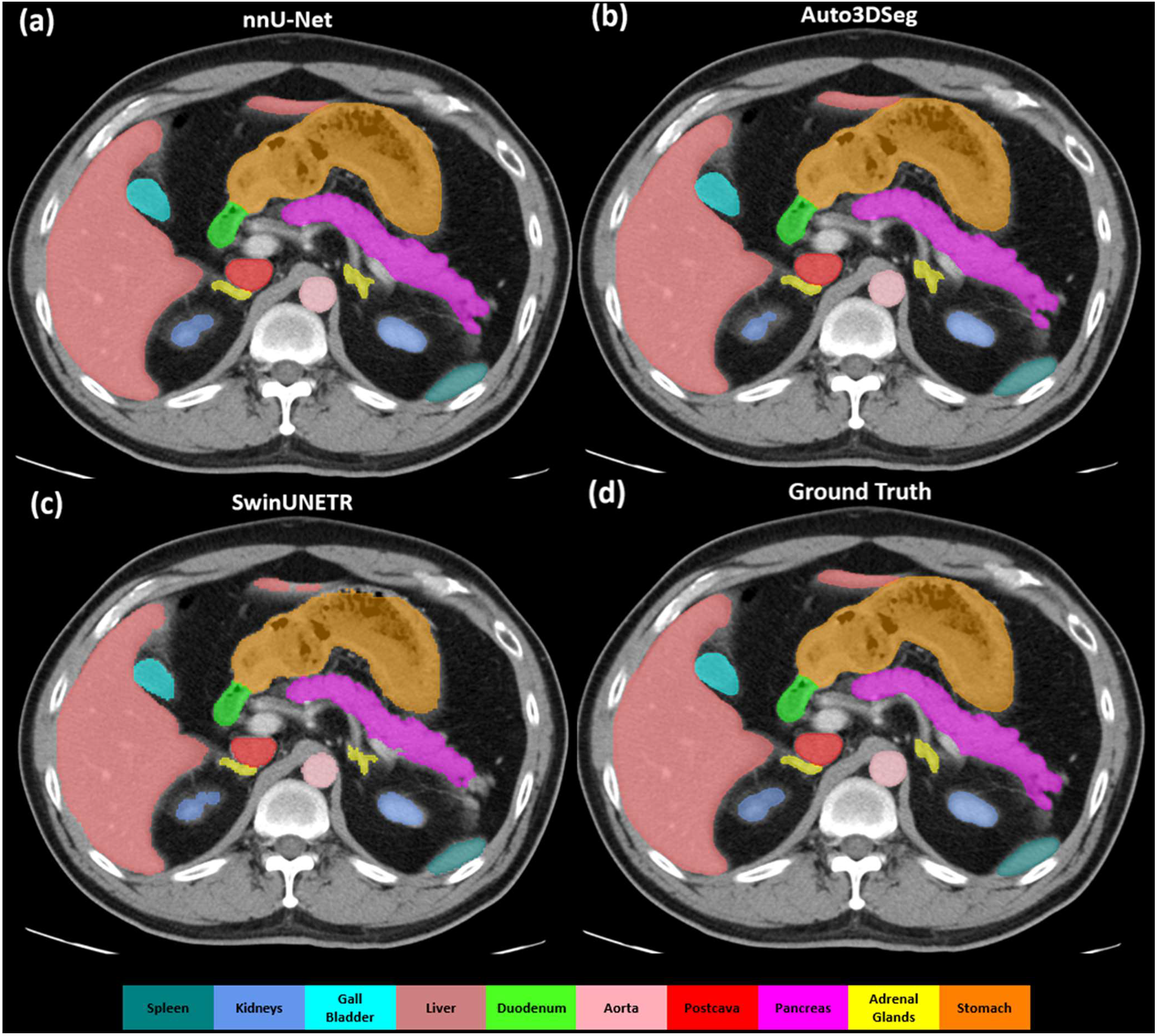
Comparisons of auto-generated (a-c) and ground-truth (d) auto-contours for individual auto-segmentation models on a holdout-validation case. (a: nnU-Net, b: Auto3DSeg, c: SwinUNETR, d: Ground Truth)

The range and median of values for all metrics are summarized in Table 3. Of particular note is the higher range of values shown by SwinUNETR in the HD95 compared to both AutoML frameworks, with Auto3DSeg showing the second highest range, and nnU-Net exhibiting the least variability of the three. Across both DSC and sDSC, the ranges for the three models are similar and expected (i.e., normalized between 0.0-1.0). Median values show a similar trend, with both AutoML models presenting higher medians than SwinUNETR. Interestingly, both Auto3DSeg and SwinUNETR present the exact same median values for the HD95, even though Auto3DSeg outperformed SwinUNETR by a large margin when analyzing mean values. This would suggest that the distributions for Auto3Dseg when it comes to DSC are positively skewed, i.e., a disproportionate number of lower scores when compared to a normal/symmetric distribution. Similarly, the distribution of the HD95 appears to be positively skewed, with a large number of lower distances. Finally, the median sDSC for nnU-Net is the highest, followed by Auto3DSeg and SwinUNETR, confirming a positive skew for both AutoML models with a lesser degree for the SwinUNETR.

**Table 3.**
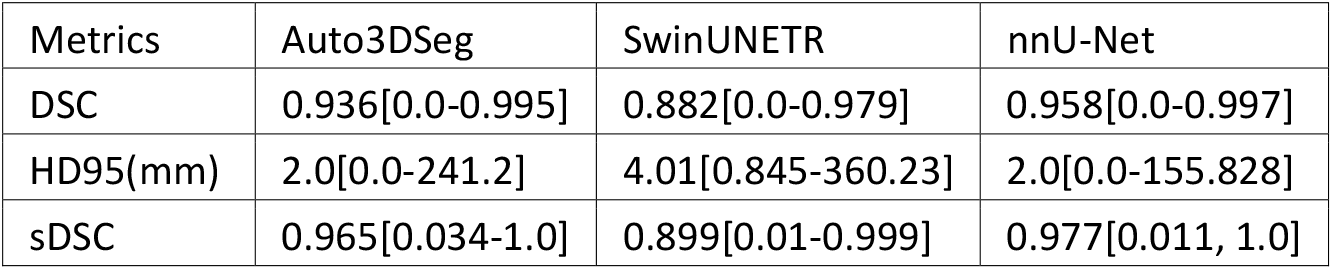
Summary of median and ranges for all OARs combined (median[min-max])

| Metrics | Auto3DSeg | SwinUNETR | nnU-Net |
| --- | --- | --- | --- |
| DSC | 0.936[0.0-0.995] | 0.882[0.0-0.979] | 0.958[0.0-0.997] |
| HD95(mm) | 2.0[0.0-241.2] | 4.01[0.845-360.23] | 2.0[0.0-155.828] |
| sDSC | 0.965[0.034-1.0] | 0.899[0.01-0.999] | 0.977[0.011, 1.0] |

### Qualitative Results

Qualitative Likert results are summarized in Figure 4, showing the performance of both AutoML models across all OARs (a, b) as well as the difference in scores across both models (c).

**Figure 4.**
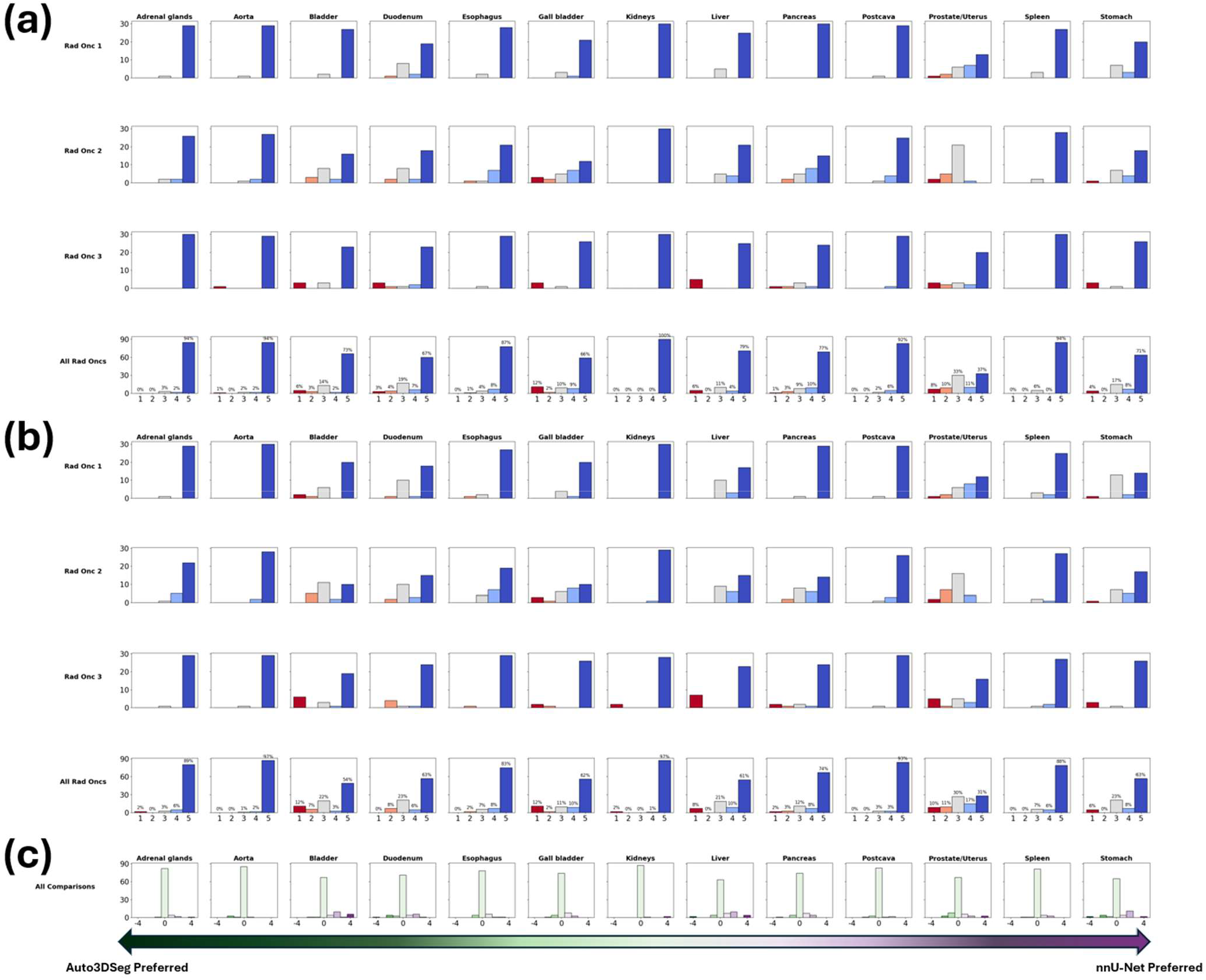
Compilation of AutoML model Likert scores per organ for all physicians, by model. (a): nnU-Net, (b): Auto3DSeg, (c): Difference of scores per organ over both models. Difference plot shows agreement between model output rankings by physicians, with positive scores indicating preference towards nnU-Net and negative scores indicating preference towards Auto3DSeg.

Considering the Likert analysis, we found that out of the 13 total OARs both frameworks contained 5 organs in common where ≥95% of contours were clinically acceptable with stylistic or no edits. These organs were: adrenal glands, aorta, kidneys, postcava and spleen. In total, Auto3DSeg had 337 contours out of 390 (86.41%) scoring a 4 or higher. Similarly, nnU-Net had 350 out of 390 (89.74%) contours scoring a 4 or higher, indicating high clinical acceptance across both models. Conducting a t-test of distributions on all 390 contours for both models resulted in a p-value of 0.0027, indicating a statistically significant preference for the nnU-Net generated auto-contours. When comparing per-organ results by way of the 2-tailed Wilcoxon signed-rank test, we found that nnU-Net was superior to Auto3Deg for the bladder and liver OARs (p-values of 0.011 and 0.047 respectively). We found no statistical significance (p > 0.05) in qualitative scoring in the remaining organs.

## Discussion

In this study, we investigated three established and widely used frameworks for auto-segmentation pipelines, SwinUNETR, nnU-Net and Auto3DSeg. To our knowledge, this is the first study performing a head-to-head comparison of auto-segmentation AutoML frameworks. We evaluated each framework using the publicly available AMOS22 dataset, acquired through the MICCAI AMOS22 grand challenge. These models were trained and evaluated both quantitatively and qualitatively.

Based on quantitative metrics, both AutoML methodologies significantly outperformed the SwinUNETR method, with nnU-Net being the highest performing of the two AutoML frameworks. DSC comparisons of both AutoML models (nnU-Net vs. Auto3DSeg) in DSC resulted in p<0.0001 for 12/13 OARs with no significance in the bladder. With HD95, there were fewer significantly different OARs, with bladder, prostate/uterus and duodenum OARs having 0.01<p≤0.001, with liver, adrenal glands and pancreas showing 0.0001<p≤0.001. The remaining 4 OARs which showed no significance were the spleen, kidneys, gall bladder and esophagus. sDSC analysis showed 3 organs (spleen, gall bladder and bladder) with no significance, and the 10 remaining OARs having 0.01<p≤0.05. Comparing SwinUNETR to nnU-Net, across all DSC, HD95 and sDSC, all 13 OARs showed statistically significant differences. Finally looking at SwinUNETR vs. Auto3DSeg, across DSC and sDSC, all 13 organs presented significantly different results. For HD95, 12/13 organs exhibited statistically significant differences, with the bladder OAR showing no significance. As highlighted by these results, the non-AutoML model was unable to perform at the same level as both AutoML frameworks on any OARs over the majority of metrics, and both AutoML models performed at a statistically significant level. For all three metrics, the nnU-Net implementation consistently and significantly outperformed the non-AutoML SwinUNETR framework, as well as the Auto3DSeg framework. Auto3DSeg also outperformed the standard SwinUNETR in all evaluated quantitative metrics.

Qualitatively, physicians showed statistical preference to the auto-contours generated by the nnU-Net framework. Auto3DSeg also performed at a clinically acceptable level, however, as highlighted in Figure 3, (c), when physicians expressed about the acceptability of model contours for a particular OAR, nnU-Net was typically preferred. nnU-Net in particular, has been shown by several studies, to be a physician preferred methodology of generating contours, when assessed qualitatively. (Kraus et al, Yu et al.), in abdominal contexts. Yu et al., in particular, showed nnU-Nets ability to generate clinically acceptable contours. Yu et al. showcased that over 90% of the auto-contours generated by an nnU-Net trained model, received a Likert score of 3 (minor edits necessary, but more time efficient to edit provided auto-contour) or greater on all organs (stomach, duodenum, large bowel, small bowel, liver, spleen, left kidney and right kidney). Similarly, Kraus et al. showed a majority of contours generated by a custom trained 3D-Unet on all investigated OARs (bladder, small bowel, sigmoid, rectum and urethra) received scores of four or higher, indicating high clinical acceptability with no edits necessary.

As highlighted by Cardenas et al.^1^, Kim et al.^29^ and many others, auto-contouring applications have the potential to save valuable clinical time and efforts. Looking more specifically at existing implementations of the nnU-Net framework, Isensee et al.^6^ proposed a modified nnU-Net pipeline specifically configured for the AMOS22 grand challenge. The trained nnU-Net model presented in this study, presented higher DSC scores of 0.924 (ours), 0.90 (Isensee et al. after post-processing, and ensembling) when compared on the task 1 dataset provided by the challenge. However, important differences that potentially contribute to this performance difference may include lateral OAR combinations, which Isensee et al. were not permitted to do due to the nature of the grand challenge submission and data provided, as well as other modifications as outlined by the study such as the multi-modal nature of the original AMOS22 Grand Challenge submission, where the present study only used single modality, CT images from the AMOS22 dataset. The mean DSC as reported by Isensee et al. on the pre-defined validation set, after an identical 5-fold cross validation was 0.914. This value was achieved by increasing batch size to 5, compared to the batch size of 2 presented in our study. Additionally, adding residual encoders as well as various experience-based configuration changes which were not implemented in the current study will have impacted the performance comparisons of both model pipelines. Due to the novelty of the Auto3DSeg framework, there are fewer examples of implementations or detailed investigations on large, open-sourced datasets. In a recent study by Myronenko et al.^30^, the authors employed the Auto3DSeg framework in a similar, abdominal segmentation setting (kidney and kidney tumor challenge, KiTs) ranking first in the Grand Challenge, with an overall DSC value of 0.835 and sDSC value of 0.723. Comparison of results indicated quantitatively better contours of kidney in this study (0.946 and 0.916 in our study for average DSC and sDSC of the kidney OAR respectively using the Auto3DSeg model presented here). It is important to note that the KiTs dataset investigated by Myronenko et al. presents a very different and particular set of data, compared to the relatively diverse and large scale of data investigated here. With the newer AMOS22 dataset, the majority of studies show performances of singular frameworks for the purpose of the grand challenge, as opposed to a large, overarching comparison of various frameworks. One of the only comparisons was conducted by the team who released the dataset, Ji et al.^16^ who benchmarked certain existing deep learning frameworks using the AMOS22 dataset, including results from a similarly trained SwinUNETR, with an average DSC score of 0.864, compared to the 0.882 reported in the current study. One of the best comparisons to the current study comes from Roy et al.^31^ who show similar performances of both the nnU-Net and a similarly trained SwinUNETR implementation with the AMOS22 dataset. Looking specifically at the DSC, both nnU-Net models demonstrate superior performance, achieving scores of 0.889 (Roy et al.) and 0.958 (ours), while both SwinUNETR implementations lag behind with scores of 0.868 (Roy et al.) and 0.882(ours). The trend persists in the evaluation using sDSC, where both nnU-Net implementations outperform SwinUNETR, securing scores of 0.917 (Roy et al., *τ* = 1*mm*) and 0.977 (ours, *τ* = 2*mm)*, compared to SwinUNETR scores of 0.892 (Roy et al., *τ* = 1*mm*) and 0.899 (ours, *τ* = 2*mm*).

Even accounting for discrepancies in tolerances used in the calculation of the sDSC between the current study and that of Roy et al, it is evident that nnU-Net consistently demonstrates higher effectiveness than SwinUNETR in abdominal segmentation tasks. Adding on to these conclusions, studies by Wan et al.^32^, and Yang et al.^33^ show that AutoML solutions often outperform the state of the art, non AutoML methods and that comparative studies such as these have the potential to translate very well clinically. The well-established nnU-Net framework exhibited commendable performance with minimal issues, proving to be a straightforward and reliable choice for medical image segmentation tasks. However, our investigation revealed a notable contrast when employing the Auto3DSeg framework, which is still in its early days of public implementation and required much more devoted time and effort before being implemented successfully. This nuance underscores the rapidly evolving landscape of automated solutions, emphasizing the need for cautious consideration of the maturity and robustness of the chosen framework.

It is also essential to acknowledge certain limitations inherent in our study. Our focus on the AMOS22 dataset, which was derived from a single institution (Longgang District People’s Hospital, Shenzhen, China), raises the need for further validation across diverse datasets to ascertain the generalizability of our results. Additionally, the reliance on a three-physician analysis from a single institution introduces potential biases in our qualitative analysis, influenced by specific institutional standards. Furthermore, the comparison of a single non-automated framework is not, in itself, a sufficient expereimental comparison to 2 SOTA AutoML frameworks, which may limit the validity of definite conclusions regarding it’s comparison. Future research endeavors should aim to address these limitations, broadening the scope of investigation to encompass a more diverse range of datasets across multiple sites and modalities as well as incorporating a multi-institutional study with several AutoML and manual frameworks for a more comprehensive understanding of AutoML’s clinical applicability in medical image segmentation.

## Conclusions

We conducted a comprehensive evaluation of 2 leading AutoML frameworks (Auto3DSeg, nnU-Net) and compared them to a state-of-the-art non-AutoML framework (SwinUNETR) in the context of abdominal OAR segmentation, using the publicly available AMOS22 dataset. We trained these models and evaluated the outputs in a qualitative and quantitative manner. We show that AutoML frameworks provide a much more time and resource efficient manner of achieving clinically acceptable results with significantly better contours than the non-AutoML framework. These findings will inform further studies into the clinical deployment of similar auto-segmentation pipelines.

## Data Availability

The data for this study was provided via the MICCAI AMOS22 Grand Challenge. (https://zenodo.org/records/7155725#.Y0OOCOxBztM)

## Acknowledgments

This work used the JETSTREAM2 high performance compute cluster at Indiana University, through allocation MED220020 from the Advanced Cyberinfrastructure Coordination Ecosystem: Services & Support (ACCESS) program, which is supported by National Science Foundation grants #2138259, #2138286, #2138307, #2137603, and #2138296.

## Author Contributions

Each author had participated sufficiently in the work to take the responsibility for appropriate portions of the content. U.R and C.E.C were responsible for the training and evaluation of the frameworks as well as the research design and amalgamation of results. J.A.P provided help with statistical analysis and manuscript review. R.J, N.T.P, and M.S offered feedback to the automatically generated contours and provided qualitative evaluations. U.R wrote the manuscript. C.E.C provided technical guidance and support for this research. All authors read and approved the final manuscript.

