## Supplementary material for "Assessing Quantitative Performance and Expert Review of Multiple Deep Learning-Based Frameworks for Computed Tomography-based Abdominal Organ Auto-Segmentation": Table 1

**Table 1.** Qualitative Scoring Criteria

| Score | Description |
| --- | --- |
| 1: (Unusable) | The automatically generated contours are so bad that they are unusable (i.e., wrong body area, outside confines of body, etc.) |
| 2: (Major edits) | Edits that the reviewer judges are required to ensure appropriate treatment and sufficiently significant that the user would prefer to start from scratch. |
| 3: (Minor edits are necessary) | Edits that the reviewer judges are clinically important, but it is more efficient to edit the automatically generated contours than start from scratch. |
| 4: (Minor edits are not necessary) | Stylistic differences, but not clinically important. The current contours are acceptable. |
| 5: (Use as-is) | Clinically acceptable, could be used for treatment without change. |
