## Supplementary material for "Assessing Quantitative Performance and Expert Review of Multiple Deep Learning-Based Frameworks for Computed Tomography-based Abdominal Organ Auto-Segmentation": Table 2

**Table 2.** Summary of quantitative metrics for all OARs combined (average +/- standard deviations)

| Metrics | Auto3DSeg | SwinUNETR | nnU-Net |
| --- | --- | --- | --- |
| DSC | 0.902 ± 0.123 | 0.837 ± 0.142 | 0.924 ± 0.116 |
| HD95(mm) | 8.762 ± 23.68 | 13.929 ± 44.85 | 4.261 ± 9.852 |
| sDSC | 0.919 ± 0.133 | 0.844 ± 0.161 | 0.938 ± 0.118 |
